# *Following the science?* Views from scientists on government advisory boards during the COVID-19 pandemic: a qualitative interview study in five European countries

**DOI:** 10.1101/2021.07.06.21260099

**Authors:** Elien Colman, Marta Wanat, Herman Goossens, Sarah Tonkin-Crine, Sibyl Anthierens

## Abstract

**Objectives:** To explore the views and experiences of scientists working on government advisory boards during the COVID-19 pandemic, with the aim to learn lessons for future pandemic management and preparedness.

**Design:** Explorative qualitative interview study.

**Participants:** Twenty one scientists with an official government advisory role during the COVID-19 pandemic in Belgium, the Netherlands, UK, Sweden or Germany.

**Methods:** Online video or telephone semi-structured interviews took place between December 2020 and April 2021. They were audio recorded and transcribed, and analyzed using a combination of inductive and deductive thematic analysis techniques.

**Results:** Scientists found working on the advisory boards during the COVID-19 pandemic to be a rewarding experience. However, they identified numerous challenges including learning to work in an interdisciplinary way, ensuring that evidence is understood and taken on board by governments, and dealing with media and public reactions. Scientists found themselves taking on new roles, the boundaries of which were not clearly defined. Consequently, they received substantial media attention and were often perceived and treated as a public figure.

**Conclusions:** Scientists working on advisory boards in European countries faced similar challenges, highlighting key lessons to be learnt. Future pandemic preparedness efforts should focus on building interdisciplinary collaboration within advisory boards; ensuring transparency in how boards operate; defining and protecting boundaries of the scientific advisor role; and supporting scientists to inform the public in the fight against disinformation, whilst dealing with potential hostile reactions.

What is already known on this topic

- To tackle the COVID-19 pandemic, governments have established various types of scientific advisory boards to provide evidence and recommendations to policy makers.
- With science becoming a focal point of this pandemic, scientific advisors also found themselves in the public eye.
- As more attention is being paid to analysing what we can do to be better prepared for the next pandemic, the views of key actors, i.e. government scientific advisors, is still largely missing.

What this study adds

- The government scientific advisors working during the COVID-19 pandemic faced a number of challenges such as working in an interdisciplinary way with their peers on scientific boards, establishing a working relationship with government allowing evidence to be taken on board, and dealing with media and public reactions.
- It is crucial that we take on board key lessons shared by scientific advisors, which calls for building interdisciplinary collaboration within advisory boards; ensuring transparency in both how boards operate and clear boundaries of scientists-government relationship; and supporting scientists in their role of informing the public.

## Introduction

Since January 2020, when the World Health Organisation has declared COVID-19 to be a global health care emergency,^1^ countries across the world have witnessed its devastating consequences. In order to tackle the pandemic, governments have established various types of scientific advisory boards to provide evidence and recommendations to policy makers.^2-4^ These scientists have been faced with numerous challenges, including working under significant time constraints to produce evidence as quickly as possible.^5, 6^

While scientists have been able to make some impact on policy since the start of the pandemic in European countries,^7-9^ the complex relationship between scientists and governments during this pandemic has been widely discussed. There have been debates about the boundaries, scope and neutrality of the role of scientific advisors, which highlighted the complexity of advisory process with some questioning whether it is even possible for scientists to provide value-free recommendations.^3, 9, 10^ There have also been reports of policy makers’ attempts to influence the scientific advisory boards, e.g. in the UK,^10^ Belgium,^11^ and the Netherlands,^12^ which highlighted at times blurred boundaries of government-scientist relationship. Some raised alarms that there has been little accountability of governments’ actions in poor handling of the pandemic,^10, 11^ highlighting powerlessness of public, healthcare professionals as well as scientific experts in challenging government’s actions^6, 10^ and called for the end of suppression of science.^10^

“Following the science” has been used by politicians as both a shorthand for a new, “better” era of politics, where government decisions will be based on scientists and public health experts advice, for example in the US,^13^ as well as an explanation or even excuse for certain government’s decisions and failings.^8^ In the context of ‘the science’ becoming a focal point of this pandemic, scientific advisors have found themselves in the public eye. While some gained a lot of recognition for their work, some have experienced personal attacks. Recently, a Belgian scientist had to move into a safe house,^14^ highlighting the extreme consequences of scientists being in the spotlight. This has been often magnified by politicization of COVID-19 with media coverage playing a key role in how science and scientists have been perceived by the public^15^ and blurred boundaries between science, policy and politics,^4^ making it difficult for public to make the distinction between scientific advice and government decision.

As more attention is being paid to analysing what we can do to be better prepared for the next pandemic,^16, 17^ the views of key actors, i.e. government scientific advisors, are still largely missing. This paper fills this gap by examining the views and experiences of scientists working on government advisory boards during the COVID-19 pandemic. The aim is to understand these experiences, to learn lessons for future pandemic preparedness, and to understand how we can better support scientists working during future health emergencies.

## Methods

### Design

This was a qualitative study using semi-structured interviews.

### Sampling and recruitment

We used purposeful and snowball sampling to recruit scientists working on advisory boards during the COVID-19 pandemic. The inclusion criteria included: i) currently working at an academic or public health (research) institution, ii) experience of an official government advisory role during the COVID-19 pandemic in any of five European countries: Belgium, the Netherlands, the UK, Sweden and Germany. These countries were chosen to provide variation in how boards operated. We aimed to recruit scientists from a range of disciplines mirroring the composition of boards across Europe.^4^ To recruit participants, three methods were used. First, as this research was part of the EU-funded Rapid European COVID-19 Emergency Response research (RECOVER) project, which aimed to inform Europe’s response to the pandemic and any future emerging infectious disease outbreaks, we recruited participants within this existing network of leading scientists. Secondly, we used snowballing sampling by asking RECOVER partners to reach out to potential participants from their own networks. Thirdly, email invitations were sent to potential participants identified from government websites where member lists of COVID-related advisory boards were available. A letter of invitation, information about the study and consent form were emailed to all potential participants. All participants gave verbal consent to take part.

### Data collection

Two female experienced post-doctoral qualitative researchers (EC and MW) conducted video or telephone interviews. The interviewers followed a topic guide exploring the meaning of being a scientist working during the pandemic; new roles and responsibilities; experiences of working with other scientists within advisory boards, collaborating with governments; and informing the public (see Supplementary File: Topic Guide). The interviews were conducted in English or Dutch, audio recorded and transcribed verbatim. Field notes were made after each interview. Interviews continued until data saturation was reached.

### Data analysis

A combination of inductive and deductive thematic analysis techniques were used.^18^ After conducting the first 10 interviews, the two interviewers read the transcripts to immerse themselves in the data. They then coded the data into 15 a priori categories based on the topic guide. Data within each category were then coded inductively line by line to create sub-categories. These were then grouped to created themes and sub-themes. These categories and sub-categories were discussed and agreed upon with the wider interdisciplinary team. This thematic framework was then used to code remaining interviews. In order to enhance the quality of the analysis, researcher triangulation and member checking was carried out;^19, 20^ this involved discussion of the data and analysis at several stages amongst the wider multidisciplinary team, comprised of psychologists, sociologists and a public health scientist. We also sent a draft of the results to all participants for their feedback. The analysis also benefited from feedback from the RECOVER scientific committee, consisting of international scientists, thus further enhancing relevance of the study findings. NVivo version 12 was used to support the analysis process.

### Patient and Public Involvement

Give that PPI was not central to the topic being explored, and a very rapid set-up of the study, PPI involvement was not possible.

## Results

Interviews with scientific advisors were undertaken between December 2020 and April 2021. The average length of the interviews was 43 minutes (range: 30 –61 minutes).

In total, 84 scientific advisors were invited to participate from five countries. Twenty one semi-structured interviews were carried out, resulting in a response rate of 25%. Three scientific advisors agreed to participate, but were not able to schedule an interview and another three experts declined their participation, after having first agreed, because of concerns related to being identified. Many participants had “out-of-office” email replies stating that they were working on scientific boards and would be unlikely to respond.

Table 1 gives an overview of the number of scientific advisors invited in each country. This varied depending on the availability of official member lists of advisory boards and the extensiveness of the network of the research team.

**Table 1.** Response rates across participating countries

|  | Belgium | Netherlands | UK | Sweden | Germany | Total |
| --- | --- | --- | --- | --- | --- | --- |
| Invited | 9 | 10 | 53 | 6 | 6 | 84 |
| Interviews conducted | 5 | 6 | 7 | 2 | 1 | 21 |

Most participants were bio-medical scientists (N=16). Five participants were social scientists. The majority was solely employed at a university (N=17), while two participants worked at a public health (research) institution and two participants combined an academic position with a position in a governmental research institution. The majority of participants were male (N=16).

We identified five themes, capturing participants’ views and experiences of working during the COVID-19 pandemic:

1. Complexities of working on scientific boards
2. Learning to present evidence and recommendations
3. Nature of the relationship between scientists and government
4. Making sense of the boundaries of their role as a scientific advisor
5. Being in the public eye

These themes are discussed below with supporting quotes.

### Theme 1: Complexities of working on scientific boards

Participants described numerous opportunities and challenges related to working on scientific boards and presenting evidence.

Participants highlighted that scientific boards were made up of scientists representing different disciplines and that the key to success of this interdisciplinary collaboration was to focus on one’s expertise and respect each other’s specialist knowledge. Scientific advisors reported a great sense of satisfaction in working with their peers, whose work they valued, and described a sense of community among scientists who enjoyed these - in many cases - new collaborations. Equally, pre-existing collaborations seemed to facilitate good working relationships. Some noted that the pandemic facilitated more collaboration across universities than pre-pandemic work, when relationships were more competitive, because participants were working towards the same goal of tackling the pandemic.

However, interdisciplinary collaboration within scientific boards did not always go smoothly, as some experts felt that not all board members valued each other’s contributions. Integrating insights from various specialisms within a single board was also a challenge when individuals felt that their area of expertise should be a priority and some participants felt that, at times, some of their colleagues overstepped their expertise, lacked openness towards other disciplines, or even questioned the expertise of their peers, which resulted in frustrations.

*[Talking about interdisciplinary collaboration within the board] That was most frustrating because we were also sitting at the table with people who didn’t want to understand. You should at least be open to the vision of one another [P19, Belgium]*

Participants felt that, at the start of the pandemic, scientific boards tended to be dominated by colleagues from bio-medical sciences, particularly virology. Social scientists in particular expressed the view that they had difficulties in making their voice heard, by not being represented to the same extent as other disciplines, on advisory boards. Related to this, participants felt that the composition of the board was very important in how “evidence was viewed”, and whether “it was accepted and taken forward” within board decisions. Participants reported that the understanding of what constitutes “evidence” seemed to differ among board members who were not always familiar with methods used by other disciplines. For example, some social scientists felt that they had to learn how to communicate their research and the methodologies used to their colleagues for their evidence to be seen as equally valid to that from other bio-medical disciplines.

*What is striking is what they consider scientific evidence […], for example, we have different types of evidence e*.*g. surveys or interviews and I really had to learn how to present this as legitimate evidence. […] I’d try to communicate in a way that people from that side would understand. […] So just to try to give some nuance to the evidence to the extent that I think I even provided more background and detail than some of the medical things that are taken immediately as truth. [P16, UK]*

However, over time, participants reported that it became apparent that a broad array of disciplines was needed to be able to inform the governments’ strategy as the pandemic had affected all aspects of society. Also, participants reflected that with time, these collaborations seemed to work better, as they learned from each other’s expertise, all having to broaden their scientific horizons, got to know each other better and built trusting relationships.

*It keeps everyone on their toes, the motivational psychologists, the economists and so on, who gradually learn the more biomedical and bio-statistical side of the story. And vice versa, for example, you learn the important motivational elements in communication, you also learn your lesson. [P14, Belgium]*

### Theme 2: Learning to present evidence and recommendations

Scientists described that working on advisory boards during the pandemic was different to how they worked pre-pandemic for a number of reasons.

First, they highlighted that different advisory boards had different ways of operating, which meant that the roles they took on varied as well. On some advisory boards, the scientists were tasked with generating and presenting evidence; on some with knowledge transfer based on their expertise in a particular field, while others involved giving recommendations or overseeing implementation of their advice. This meant that they often had to switch roles and the ways in which they worked.

*So for example on X [name of scientific committee], the data and our opinions would be collected and we would have to say: ‘How certain are we of this opinion?’ So we would say: ‘Low certainty, medium certainty, high certainty*.*’ So for the Y [name of scientific committee], that was more to try to help ministers understand some of the concepts. And also, to say why we didn’t know […]. So there were different types of functions. [P10, UK]*

Secondly, participants were often tasked with providing evidence and/or recommendations very rapidly. They highlighted that the ever-changing pandemic situation meant that they were trying to gain a quick understanding of the emerging evidence, which was also constantly changing. This meant having to work very long hours to meet tight deadlines, in order to provide evidence before it became outdated.

*If you have results that are two weeks old, they are already a bit outdated and of limited use. And that is of course a completely different kind of speed […] Researchers can be perfectionists and during a pandemic you really need to find the balance between what are the most important things that you really want to get right (I*.*e. quality), and what are those things you can deliver within those few weeks (I*.*e. speed). [P21, the Netherlands]*

*You actually have to think very quickly, react very quickly and also develop your opinion very quickly, which of course contrasts with the slowness of science […] I constantly get pushed out of my comfort zone. [P15, Belgium]*

Experts also faced difficulties when being tasked with making recommendations when evidence was limited, inconclusive or not available. They highlighted that the pre-pandemic way of providing evidence involved feedback from colleagues, e.g. peer review, and presenting it in a more nuanced way. In contrast, they were now tasked with providing an answer to specific questions and offering a more definitive answer, which challenged their previous ways of working. This also meant that at times, public pressure affected recommendations, by forcing scientists to still make a recommendation, despite limited or inconclusive evidence.

*We have had endless discussions about those face masks on the street and our opinion was that you really have very little evidence for that. But there was a lot of social pressure, so advice was actually drawn up under social pressure*. [P4, the Netherlands]

Because of this lack of evidence, participants felt that the recommendations were sometimes susceptible to questioning by other experts, opinion makers, government members or other stakeholders.

*You work in such a crisis with a very high degree of uncertainty. And that is very difficult, isn’t it, because then you often get stories such as the fitness sector says “yes, but where is the proof that we contributed to the increase in that second wave? Well, you just can’t give that proof because you just can’t get that data out of the system. [P8, Belgium]*

### Theme 3: Nature of the relationship between scientists and government

Scientists highlighted a number of barriers and facilitators to a good relationship between advisory boards and government. Overall, scientific advisors felt that collaborations were somewhat working and that, in comparison to both how and the extent to which experts were consulted pre-pandemic, scientists had a more profound impact on policy decisions during the COVID-19 pandemic, which they found rewarding.

*You very quickly see the benefits of generating knowledge and taking it through to policy very quickly. […] I’m collecting possible data, putting it together and generating reports and seeing that go straight to policy. So it’s totally different from what we were doing in the pre-pandemic phase. [P9, UK]*

Nevertheless, they also described tensions between advisory boards and policy makers and mentioned two main challenges. Firstly, they highlighted that governments did not always take on board their evidence or did not take it on in a timely manner. Secondly, some participants felt that at times governments presented policy decisions as if they were solely based on scientific evidence, which they thought was misleading for the public. Some scientific advisors also felt that politicians were “using” the advisors to justify political decisions in their communication to the public, especially related to unpopular restrictions.

*At some point in March, our Prime Minister said “I do what the [name of advisory board] says”. I found that a very unpleasant comment, because it implies that you are directly responsible for the policy pursued and that is not the case. It is simply a political choice. And we only give advice. So they should take it as such and not hide behind that advice. [P4, the Netherlands]*.

However, similarly to their relationships with their peers on the boards, scientific advisors felt that over time, collaborations had improved. They attributed this to policy makers gaining a better understanding of scientific methods and how evidence was generated and improved communication. Some experts also described that they learned how to formulate their recommendations more clearly and in a more implementable and compelling way, which improved the chances of it being used by policy makers. For example, using GRADE criteria to present evidence was considered a transparent and practical way of presenting evidence.^21^

### Theme 4: Making sense of the boundaries of their role as a scientific advisor

Scientists tried to make sense of what their role as a scientific advisor was and what the boundaries were. This seemed to be not only linked to how advisory boards operate (as described in theme 1), but also how they felt about their role. That meant that participants interpreted their role in different ways and differed in how they saw their relationship with governments.

As described in Theme 1, scientific advisors were at times faced with situations when the evidence was not translated into policy. They expressed a variety of views in relation to how they felt about it and whether they had taken any action to “rectify” this issue. Most scientific advisors expressed frustration and disappointment when they had faced such situations especially when they did not receive any feedback on the decisions made in relation to (not) implementing certain measures. Despite this, most participants viewed their role as “just” providing evidence and recommendations to policy makers, and felt that it was the responsibility of the government to make decisions and implement them.

*And the politicians have to make the final decision because they see the whole picture. We must be very careful as experts not to become politicians and not to cross that line. [P2, Belgium]*

*I sometimes am surprised by the decisions that are being made by the politicians but, ultimately, it’s them who decide. […] We sometimes express a very strong view that, from the scientific point of view, our consensus is absolutely clear that we now have to introduce various lockdown measures and they just don’t’ happen. But I do understand that the government has a range of pressures upon them and that our scientific view is just one of them. […] They’re elected representatives and I’m not there to say that they’re wrong, but just to say what the point of view of scientists is. [P3, UK]*

Consequently, many were reluctant to take an active stand when their advice was not taken on board in their official capacity as advisors. They described their reluctance to comment on political decisions in the media as they wished to maintain a good working relationship with policy makers. They wanted to stand apart from political arguments and to assert their scientific advisory role. Also, they were reluctant to question policy decisions publicly, as they did not want to undermine the public trust in the measures that were implemented.

*Well, I am generally always very cautious about criticizing the policy pursued. Because I think that does not contribute to the support in society. There is already so much disagreement about how to handle that Corona crisis. [P20, the Netherlands]*

Some also highlighted the dangers of scientists taking a standpoint and the consequences of universities or public institutions appearing to be lobbying for certain policies to be implemented. In addition, some scientific advisors stated that, at times, certain politicians had asked them to withhold from commenting on policy decisions. Notwithstanding these reasons to hold back from publicly commenting on policy decisions, participants also valued their independence and academic freedom. Consequently, in some cases, experts did not simply accept policy decisions. They actively expressed their concerns in the media and tried to highlight the scientific evidence on the contested issue in order to achieve some kind of equilibrium between protecting and using their academic freedom, while maintaining a constructive attitude towards the government, which was not always easy.

*That is an equilibrium that you have to maintain: on the one hand, remain critical and have the feeling that you are able to express your thoughts as a scientist, but at the same time, you cannot be diametrically opposed to politicians. [P8, Belgium]*

*We have always said: we must be able to continue to communicate from a position of academic freedom. That is important. It has also made that your words weigh more heavily when you are a member of a board, and that is something you have to learn to take into account. [P14, Belgium]*

Scientific advisors felt more inclined to influence policy makers through the media in cases when the channels of communication between scientists and governments were not perceived to be working and their recommendations concerning some crucial topics were not turned into policy. In these situations, they felt that media outlets could offer a solution to make sure that the public received the information they deemed necessary and to influence policy makers indirectly, by making their voice heard. Experts often highlighted that in those occasions they had taken the opportunity to actively have their voices heard but making it clear that they are speaking in a personal capacity rather than on behalf of a scientific board.

*There have been fewer channels to communicate between researchers and policy makers. And I think that could be one of the explanations why researchers have felt maybe obliged or forced to discuss their views in the public eye, in the media, rather than having a direct channel to policy makers where one could have a much more nuanced discussion with pros and cons. [P11, Sweden]*

*I wrote an article […]. Again, to bring this to public awareness. Again, to try and influence policy and get schools to reopen, and again just to bring it to public awareness, my concern about the ongoing psychological impact on mental health and well-being. So yeah, again it was just trying to get people to think, to engage, and again, to bring [my field] to the public awareness [P18, UK]*

### Theme 5: Finding oneself in the public eye

Participants reflected on their motivations for joining advisory boards. They felt a sense of responsibility to use their expertise and be part of the scientific community helping to tackle the COVID-19 pandemic, which felt rewarding.

Participants’ motivations were to some extent reflected in how they viewed their role of communicating with the public. They described taking part in media interviews with the aim to ensure that the public was informed of new developments and scientific insights. Especially at the beginning of the pandemic, participants considered educating and informing the public about the COVID-19 virus and the pandemic as a key part of their role as a scientist.

*I’ve always enjoyed trying to explain things to people in simple terms. […] I think in general it has been remarkably positive. The reason is because people are grateful for any clear understanding they can have of what a virus is. [P12, UK]*

*Particularly in the first half, there was a great need for information. People didn’t really understand what was happening. And people wanted explanations. […] I thought it was great that I could contribute to that. That I, that I could give that explanation to people* [P20, the Netherlands]

Multiple experts highlighted that, over time, the content of their media contributions shifted from explaining scientific insights towards explaining the measures that were taken to tackle the pandemic. Participants felt that it was important to have a meaningful role in public engagement to ensure that the public had access to correct evidence-based information, to tackle ‘fake news’ and disinformation.

*This has taken a lot of time, but I also felt like that is something that I wanted to prioritize, because I think it’s important to try to explain the trends to people. I see so many weird, strange discussions and debates. So, it sort of feels like it’s important to contribute to that discussion to make it more evidence based. I feel I have a role to play that could be important. [P11, Sweden]*

However, as a result, many experts found themselves in the public eye and consequently, became very well-known figures. Developing a relationship with the media was one of the main challenges, with participants describing both successful and less successful examples. Some journalists were keen to hear from scientists and give them a voice and space to talk to the public; participants highlighted that some journalists worked very hard to make sure that science was described accurately, by working closely together with scientists.

*I usually think they do that very neatly, they always show quotes from me in advance, to see if the content is correct. And if I want, I can always adjust it*. [P5, the Netherlands]

However, participants also experienced less successful examples of media relations where media organisations played different scientists off against each other or against policy makers (sometimes without prior warning); used quotes taken out of context; called them unexpectedly and pressured them into providing a more black and white picture of the situation than they were prepared to present. Many highlighted that engaging with media took a lot of time, as the media attention was excessive, and they wanted to make sure that they were prepared for interviews.

Some participants highlighted that the press created a particular narrative around government decisions linking individual scientists to certain political decisions, as did some politicians (as described in the Theme 2). Because of this, certain scientists were perceived by the public as responsible for government decisions.

*The perception of responsibility is entirely wrong, because political decisions always take science as one of their inputs, while sometimes the public perceives political decisions to be entirely destined by scientific information, which is just not the case. […] Because of the picture that the media creates, it’s really a narrative in the media. [P7, Germany]*

As a result of their exposure in the media scientists faced a variety of reactions from the public. While many described positive responses from the public, which included getting “thank-you” messages, others also described receiving numerous negative messages, including death threats.

*I have gotten a lot of death threats. So I get them emailed to me but I even get people calling work, you know agitated trying to find me. It’s quite scary actually so I had to make police report. [P16, UK]*

Some participants felt that there was limited support available to them to deal with such threats as it was not clear whether it was the role of an employer or solely the responsibility of the police to deal with such issues, while others felt that their negative experiences had been handled well.

Despite these negative reactions, many felt that it was important to continue engagement with the public.

*I believe we have to go on the media to put the point of the scientists across because unfortunately, there are things on social media that are just fake news and disinformation. […] I think it’s the duty of scientists to put the truth across as far as we know it. So I think we just have to accept that this is one of the downsides. [P10, UK]*

Some scientists also highlighted that presenting scientific evidence is a skill and that the public should be made aware that science evolves. While the current scientific evidence points in one direction, future research can challenge previous assumptions and scientists need to adapt their thinking and communicate these developments to the public.

*You really need to understand and live with the knowledge that this is constantly changing. You can never be quite sure that what science delivers to you one month, or the next month. And you need to be transparent about that when you communicate about it. [P6, Sweden]*

A number of participants also highlighted the importance of transparency around the content of recommendations made by advisory boards, which could be facilitated by publishing the minutes of the meetings. This would help highlight that scientific evidence was usually much more differential than the black and white picture of the situation often presented in the media or portrayed by politicians. Participants emphasized that differences between advice delivered by advisory boards and policy decisions made by governments should be clear.

## Discussion

### Principal findings

Our study showed that scientific advisors found the experience of working during the pandemic rewarding but that they also faced numerous challenges. These included learning to work in an interdisciplinary way and respecting each other’s expertise, ensuring that evidence is understood and taken on board by governments, and dealing with media and communication with public. Scientists found themselves taking on new roles, the attributes of which were not clearly defined, and thus were interpreted in various ways. Because of these new roles, the scientists received much media attention and were often perceived and treated as a public figure. This study highlights several lessons which can facilitate preparedness for future health emergencies.

### Strengths and weaknesses of the study

To our knowledge, this is the first international study exploring the experiences of scientists working on government advisory boards during the COVID-19 pandemic, thus giving a voice to key actors in this pandemic. The heterogeneity of our sample, who consisted of both medical and social scientists in five European countries, has enabled us to examine a variety of perspectives. We also note some limitations. Firstly, the limited number of participants from some countries prevented us from making cross-country comparisons. It was encouraging though that scientists’ views within and between countries have largely been consistent, highlighting that the key tensions and opportunities of working as a scientist during the COVID-19 pandemic were shared between contexts. Secondly, the current study only describes the views and experiences of scientists advising policy makers at a national level. Exploring views of scientists working on international scientific boards might also be beneficial. In addition, giving a voice to policymakers could also lead to valuable lessons on how to improve the collaboration between scientists and policy makers. Thirdly, interviews with scientists were conducted during the second and/or third waves in their respective countries. Interviewing scientists during the first wave might have shed a different light on their experiences; however, by asking them to reflect on what has changed, we were able to capture some of their views of working during the initial stages of the pandemic retrospectively. Finally, it is important to acknowledge the role of the research team in relation to the topic of study; as the researchers conducting the interviews were scientists themselves, who on a smaller scale also responded to the COVID-19 pandemic, they have regularly reflected on their own perceptions, and experiences of working during the pandemic, in order to “put these views aside” and ensure the accurate representation of participants’ voices.

### Comparison with other studies

Our study brought out issues related to the cross-disciplinary and interdisciplinary collaboration within scientific advisory boards, which was not always successful, especially at the start of the pandemic. Over time, board members learned to value each other’s expertise, integrating the insights and diverse types of evidence from the various specialisms. Others also highlighted that what counts as a “fact” differs across disciplines including what we understand as evidence, and what counts as reliable data and methods of data collection.^22^ While some scientific boards existed pre-pandemic, they operated on a much smaller scale with a limited number of scientists and some were established during the pandemic.^4^ Establishing new relationships takes some time and the limited pre-existing interdisciplinary ties posed a challenge within scientific boards in which a wide spectrum of specialisms were represented.

Secondly, our study also highlighted that the role of scientific advisors was somewhat unclear. This was linked to both the way boards operated but also how, particularly at the start of the pandemic, scientists were portrayed by both media and policy makers as responsible for policy decisions. As also highlighted by others,^3, 9^ participants described that at times, their recommendations were positioned by governments as key or the sole basis for decisions and felt that policy makers used experts to justify their decisions. For policy makers, aligning with science is considered an effective risk communication strategy.^23^ It becomes problematic, however, when decision making is clearly not in line with recommendations made by the scientific advisory boards, which has occurred on various occasions, as described by the participants in this study and others.^9^ Within this confusing context, scientists in our study highlighted that they had to establish and make sense of their role and in fact had varied interpretations of it. While the majority felt they should withhold from publicly commenting on policy decisions, some felt that they should speak up when key recommendations were not taken on board. While scientists highly valued their independence, they also restrained themselves from commenting on policy decisions, which at times was motivated by the wish to maintain a good relationship with decision makers as they had to continue the collaboration. Moreover, at times, our participants highlighted that policy makers have pressured them to not discuss certain issues in the media, while media has often urged them to comment on policy decisions.

Thirdly, scientific advisors received substantial media attention and found themselves in the public eye, sometimes unwillingly. This was a result of a variety of factors, including speaking up when recommendations were not taken on board and being portrayed as being responsible for policy decisions (as discussed above), but also because of their endeavors to inform the public in the media.

Finally, scientists reported that evidence on COVID-19 has frequently shifted, and uncertainties remained, making producing and communicating sound advice and recommendations challenging. Moreover, given the urgency, scientists had limited time to provide evidence and recommendations, which did not allow them to undergo the traditional scientific processes to guarantee and improve the quality of their contributions, e.g. peer review. Others also highlighted that the gaps in evidence can lead to diverse interpretations among scientists, policy makers and the public, thus making value-based judgements inevitable.^9^

### Lessons learnt and policy implications

Our study identified a number of lessons which can facilitate preparedness for future health emergencies. These are summarized in Table 2.

**Table 2.** Summary of recommendations in relation to key issues

| Key issues | Recommendations |
| --- | --- |
| **Need for interdisciplinary collaboration** | • Provide and strengthen entry points for all relevant disciplines in responding to the pandemic or health emergency- bringing added value to clinically and bio-medically oriented work<br>• Provide training and promote development of skills facilitating interdisciplinary working as part of professional development |
| **Importance of clear role boundaries for scientists on government advisory boards** | • Provide clear definitions of advisory and decision-making roles, alongside a transparent roadmap of responsibilities, tasks, and procedures in order to facilitate and enhance effective and trustworthy science advisory process |
| **Dealing with emerging and changing “evidence” while providing recommendations** | • Promote clear process of assessing the strength of available evidence<br>• Communicate transparently scientific uncertainties and be explicit in order to maintain trustworthiness and facilitate public trust in science<br>• Identify strategies to tackle mis- and dis-information |
| **Supporting scientists in communicating with public** | • Communicate current scientific understanding in an approachable, clear and transparent way, separate from political communication |

Firstly, our findings highlight that further development of scientists’ skills is needed to promote interdisciplinary working. For example, in the initial stages of the pandemic, social and behavioural sciences were reported not to be given centre-stage as governments focused on understanding the nature and epidemiology of the virus.^25^ However, social and behavioural sciences could have important contributions related to questions around social practices related to transmission of the virus, disease perception, or help seeking behavior as has been shown in other outbreaks as well.^26^ Looking for entry points where other disciplines can bring added value to some of the more clinical or biomedically oriented work can improve advisory boards’ preparedness to perform their expert roles during future crises.^27^ As measures to tackle a pandemic touch every aspect of society, it is key to involve experts of a wide range of disciplines who have the skills to work in an interdisciplinary fashion.^4, 28^ Secondly, this study has highlighted a need to better clarify the role of scientific advisors towards policy makers, media and the broad public. Clear definitions of advisory and decision-making roles, as well as a clear roadmap of responsibilities, tasks, and procedures in protecting the boundaries of the expert role and to facilitate an effective and trustworthy science advisory process.^4, 6, 28^ Thirdly, our study showed that scientists at the heart of the fight against COVID-19 invested time in regular and social media contact to disseminate accurate and timely information. In doing so they hoped to help tackled mis- and disinformation, which has already been named an “infodemic”.^29-31^ However, facilitating public understanding of science is also an important task for public health institutions and governments^32, 33^ and more research is needed to identify effective strategies to tackle mis-and disinformation.^32^ Finally, our study highlighted the need for scientific uncertainties to be explicitly assessed and communicated transparently to maintain trustworthiness and facilitate public trust in science.^4, 28, 34^ The need for scientists to learn to communicate science in an approachable, clear and transparent way^31^ as factual, transparent communication, which is separated from political communication, is key during a crisis.^32^

## Conclusions

Since January 2020, scientists have played a key role in tackling the COVID-19 pandemic. Following the science (or not) has become a shortcut for how this health crisis and response to it, has been perceived, used and presented by the public and governments. While many countries have started to evaluate their COVID-19 response, it is crucial that we take on board key lessons shared by scientific advisors, which call for building interdisciplinary collaboration within advisory boards; ensuring transparency in both how boards operate and define and protect the boundaries of the scientists-government relationship; and supporting scientists in their role to inform the public. While there may not be easy solutions to all these issues, countries may learn from each other’s experiences in how to increase transparency in their advisory and decision-making processes and protect the trustworthiness of science and scientific advisors.

## Data Availability

Participant level data cannot be shared without approval from data custodians owing to local information governance and data protection regulations.

## Ethics approval

The study has received ethical approval from the Ethics Committee of Antwerp University Hospital (20/13/150).

## Transparency statement

The manuscript’s guarantor (SA) affirms that this manuscript is an honest, accurate, and transparent account of the study being reported; that no important aspects of the study have been omitted; and that any discrepancies from the study as originally planned have been explained.

## Dissemination to participants and related patient and public communities

We will disseminate a lay summary of our findings through our communication channels. The involved universities will lead on the dissemination of our results to the lay audience, with help from our communications and outreach teams.

## Contributorship statement

MW and EC designed the study, co-lead data collection, co-lead the data analysis and co-wrote the first draft of the manuscript. HG secured funding, and helped conceptualize and design the study and discussion of the findings. STC and SA secured the funding, conceptualized and designed the study, and oversaw data analysis. SA is the guarantor. All the authors critically revised the manuscript drafts and approved the submission. The corresponding author attests that all listed authors meet authorship criteria and that no others meeting the criteria have been omitted.

